# Dual-energy computed tomography imaging with megavoltage and kilovoltage x-ray spectra

**DOI:** 10.1101/2023.06.22.23291766

**Authors:** Giavanna Jadick, Geneva Schlafly, Patrick J. La Rivière

## Abstract

**Purpose:** Single-energy computed tomography (CT) often suffers from poor contrast, yet it remains critical for effective radiotherapy treatment. Modern therapy systems are often equipped with both megavoltage (MV) and kilovoltage (kV) x-ray sources and thus already possess the hardware needed for dual-energy (DE) CT. There exists an unexplored potential for enhanced image contrast using MV-kV DE-CT in radiotherapy contexts.

**Approach:** A toy model comprising a single-line integral through a two-material object was designed for computing basis material signal-to-noise ratio (SNR) using estimation theory. Five dose-matched spectra (three kV, two MV) and three variables were considered: spectral combination, spectral dose allocation, and object material composition. The single-line model was extended to a simulated fan-beam CT acquisition of an anthropomorphic phantom with and without a metal implant. Basis material sinograms were computed and synthesized into virtual monoenergetic images (VMIs). MV-kV and kV-kV VMIs were compared with single-energy images.

**Results:** The 80kV-140kV pair typically yielded the best SNRs, but for bone thicknesses greater than 8 cm, the detunedMV-80kV pair surpassed it. Peak MV-kV SNR was achieved with approximately 90% dose allocated to the MV spectrum. For the CT simulations, MV-kV VMIs yielded a higher contrast-to-noise ratio (CNR) than single-energy CT at specific monoenergies. With the metal implant, MV-kV produced a higher maximum CNR and lower minimum root-mean-square-error than kV-kV.

**Conclusions:** This work quantitatively analyzes MV-kV DE-CT imaging and assesses its potential advantages. This technique may yield improved contrast and accuracy relative to dose-matched single-energy CT or kV-kV DE-CT, depending on object composition.

## 1 Introduction

When imaging for radiation therapy, high soft tissue contrast is essential. Tumors must be accurately imaged at all stages: prior to, during, and after treatment, for dose calculations, patient positioning, and cancer monitoring, respectively.^1^ The current imaging standard in radiation theory is single-energy (SE) computed tomography (CT), which often suffers from poor tissue contrast. In diagnostic imaging, dual-energy (DE) CT is routinely applied to enhance contrast. By acquiring multiple spectral data points, sufficient information is available for the calculation of basis material images and virtual monoenergetic images (VMIs). With carefully chosen contrast agents and x-ray spectra, image quality may be greatly improved using DE-CT.^2–5^

To achieve multiple simultaneous CT acquisitions, DE-CT imaging requires special hardware such as multiple source-detector arrays, energy-discriminating detectors, or fast kV-switching sources. A system lacking this hardware may also perform DE-CT by means of repeat acquisitions, but this method can suffer from misalignment or motion artifacts. Modern radiation therapy treatment systems are often already equipped with dual x-ray sources: a megavoltage (MV) source for treatment and a kilovoltage (kV) source for on-board imaging. Given this readily available equipment, there exists an unexplored potential for enhancing image quality in the context of radiation therapy using MV-kV DE-CT.^5, 6^

There are several reasons to search for new methods to improve contrast in radiation therapy settings. CT is currently the only accepted imaging modality for three-dimensional dose calculations, as it provides empirical information on electron density and atomic composition. Yet, CT suffers from low soft tissue contrast, a lack of functional information, and in some cases metal artifacts. To overcome these drawbacks, there has been recent interest in magnetic resonance (MR) and positron emission tomography (PET) imaging at different stages of the treatment planning process.^1, 7^ MR- and PET-linac systems are currently being developed and even introduced in some clinics.^8, 9^ Though these systems show promise, they are emerging and expensive, and CT imaging still remains necessary for dose calculation. Combined MV-kV imaging has the unique advantage of utilizing existing hardware, resulting in a much lower barrier to implementation and the potential to be realized in a shorter time frame. Moreover, MV images have the potential to be acquired during patient treatment, providing dual energy information to augment kV images without necessitating additional dose.^6, 10, 11^

While some prior work has explored ways to combine MV and kV information for various applications, work specifically exploring MV-kV DE-CT is limited. This may be due to the drawbacks of MV images, which can potentially contaminate kV images and reduce image quality if the two are combined naively. Since MeV photons are generally more penetrating than keV photons and have greater dose deposition per photon, MV images typically have lower contrast and greater noise relative to kV images with the same dose.^11, 12^ There are limited situations in which MV imaging alone is sufficient. For example, MV localizers or CT images have been shown to be sufficient for radiotherapy setup verification, which is useful for linacs lacking a kV x-ray source.^13^

When strategically implemented, MV information can be combined with kV information to achieve better soft tissue contrast than could be achieved with either source alone. Various techniques have been described previously.^5, 11, 12, 14–20^ For example, the greater penetrability of MV photons becomes an advantage when imaging highly attenuating objects; thus, MV data may be synthesized with kV images for metal artifact correction.^17, 20^ Similar methods have been implemented for target tracking during radiotherapy.^15, 16^

Previous work has shown promise for MV-kV DE-CT.^5, 11, 12, 14, 18, 19^ With only partial angular information, combined MV-kV CT images can achieve superior image quality relative to kV or MV alone by inheriting the advantages of each single-energy image: higher contrast from kV images and reducion in streak artifacts from MV images. These techniques utilize 90-110*^◦^* of data from each spectrum, possibly with a small amount (10-15*^◦^*) of overlap, then implement either a linear gray scale conversion or histogram mapping of pixels to reconstruct a single kV or MV image with complete angular information. Though these methods improve efficiency by reducing rotation time, the imperfect spectral mapping can cause artifacts and reduce image quality. To our knowledge, previous work focusing specifically on MV-kV DE-CT has not considered datasets with complete angular information in both MV and kV domains.

The precise conditions for MV-kV DE-CT to yield improved image quality have not been fully characterized. The experimental nature of this past work, utilizing image quality phantoms, limits the number of data points feasible for robust analysis and optimization of MV-kV DE-CT. These methods typically acknowledge the drawback of greater dose deposition by MeV photons but do not consider the effect of dose distribution between MV and kV spectra. They observe that image quality is best when MV beams penetrate more highly attenuating material inserts, but they image only a small number of material inserts within image quality phantoms. A more complete analysis should assess image quality for a continuum of material attenuations and spectral dose distributions.

In this work, we implement the analytical method proposed by Roessl et al., which uses estimation theory to characterize basis material signal-to-noise ratio (SNR) along a single-line integral with dual-energy x-rays.^21^ This method has not yet been applied to MV-kV imaging. With this technique, we are able to survey a wide range of parameters without experimental measurements, facilitating more robust characterization of situations in which MV-kV imaging provides superior image quality and quantification of the degree of improvement. We consider three variables: spectral combination, dose allocation between the two spectra, and material composition. To gauge utility of MV-kV DE-CT in a more clinically realistic setup, we also extend this model to a CT raytracing simulation with parameters informed by the single-line optimization.

## 2 Methods

Two models were developed for quantifying basis material image quality: a toy model using a single-line integral with a two-material object and a fan-beam CT simulation with a computational anthropomorphic phantom. The single-line model was used to maximize basis material SNR as a function of spectral dose allocation for an object with various bone thicknesses. These results were used to inform the simulated CT imaging task.

### 2.1 Signal detection framework

The detected signal *λ* with each spectrum *i* was calculated as

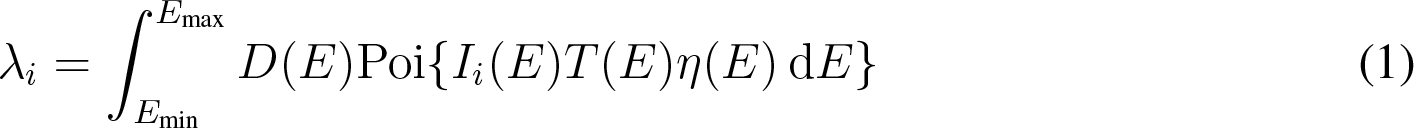

where *E* is the energy, *I_i_*(*E*) is the incident x-ray spectrum (photons per energy), *T* (*E*) is the object transmission function, *η*(*E*) is the detective efficiency function, *D*(*E*) is the detector response function, and the notation Poi{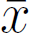} indicates generation of a realization of a Poisson random variable with mean 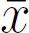. In Eq. 1, the argument of the Poisson noise is the mean number of photons of energy *E* stopped by the detector. This indicates that the signal noise model is compound Poissonian weighted by *D*(*E*).

Transmission *T* (*E*) was computed as the line integral attenuation through the object of interest,

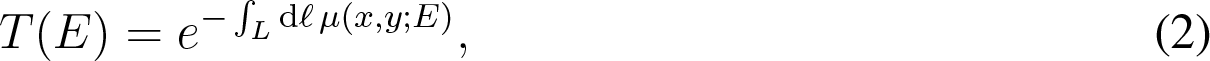

where *ℓ* is the distance along the given ray *L* and *µ*(*x, y*; *E*) is the linear attenuation coefficient of the material at location (*x, y*) evaluated at energy *E*.

An energy-integrating detector was modeled (*D*(*E*) = *E*) with detective efficiency *η*(*E*) as shown in Fig. 1. The detective efficiency function was computed to yield performance consistent with that of a previously described high-DQE detector for megavoltage imaging.^22^

**Fig 1.**
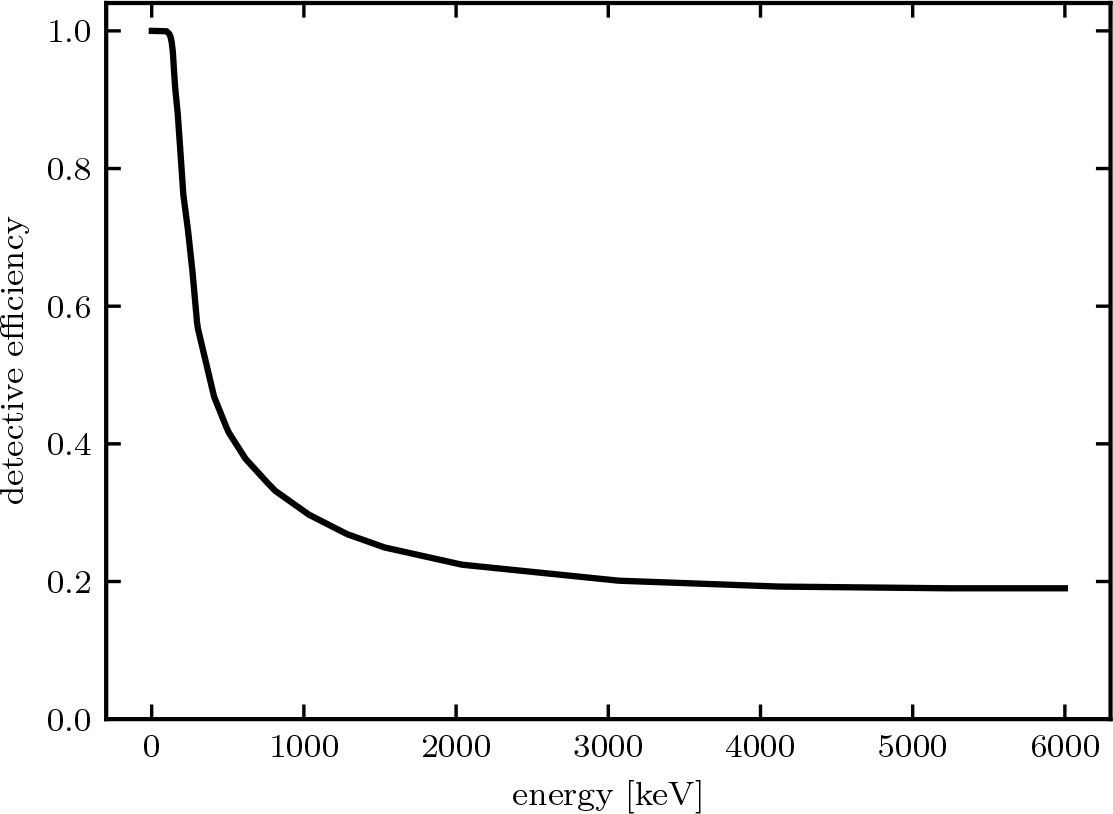
The modeled detective efficiency function *η*(*E*).

#### 2.1.1 Input spectra

Five polychromatic spectra *I_i_*(*E*) were modeled (Fig. 2). Three kV spectra were chosen to represent common diagnostic CT options (80kV, 120kV, and 140kV). The two MV spectra were modeled after a typical treatment beam (6MV) and a treatment beam with energy detuned to below 3 MV for imaging (detunedMV).^23, 24^ To ensure a valid basis for comparison, the flux of each spectrum was scaled to deliver the same dose to the center of a 40-cm diameter water cylinder (depth *d*_w_ = 20 cm) under the condition of charged particle equilibrium,

**Fig 2.**
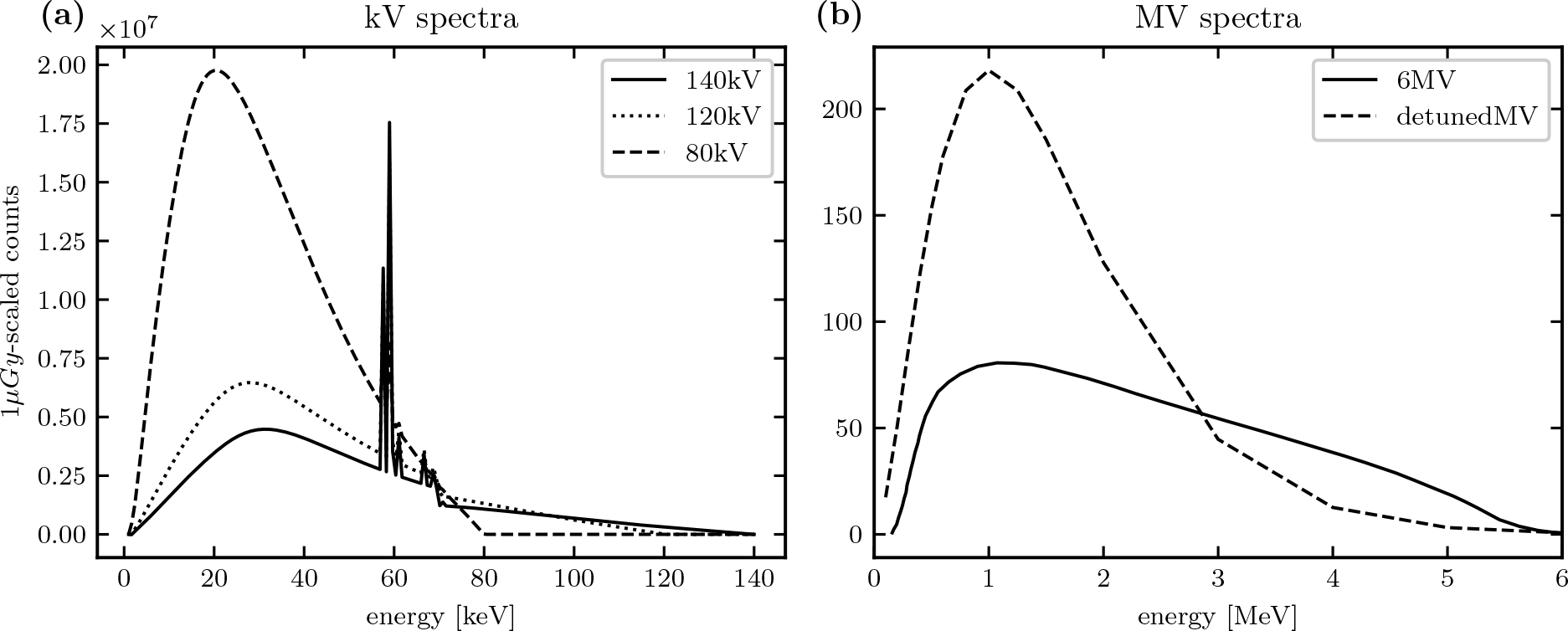
The five spectra with magnitude scaled to deliver 1 µGy dose.

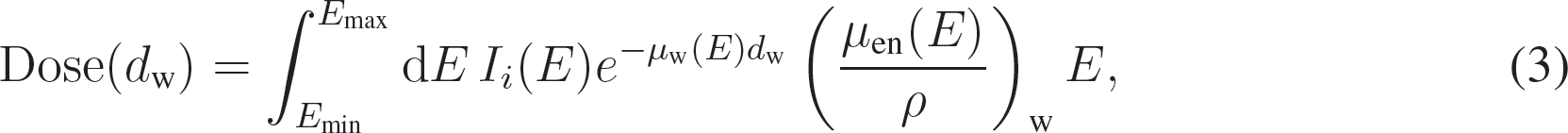

where *µ*_w_(*E*) is the linear attenuation coefficient of water and (*µ*_en_(*E*)*/ρ*)_w_ is the mass energy absorption coefficient of water.^25^

### 2.2 Model 1: single-line integral through a two-material object

Basis material SNR was computed using an estimation theory framework for a single ray incident on a two-material object of ICRU tissue and bone with densities *ρ*_tissue_ = 1.00 g/cm^3^ and *ρ*_bone_ = 1.85 g/cm^3^.^21, 26^ The tissue thickness was fixed at *t*_tissue_ = 40 cm and the bone thickness was varied from *t*_bone_ = 1 to 10 cm. All MV-kV and kV-kV spectral pairs were considered, yielding nine DE combinations. The total single-line dose allocated to both spectra was set to 1 µGy. For a typical CT acquisition with near 1000 projection views, this would sum to a dose of 1 mGy. SNR was characterized as a function of dose allocation *r* in 1% increments (from 1% to 99%), where the high-energy spectrum was rescaled by *r* and the low-energy spectrum by 1 *− r*.

The SNR for each basis material *j* was defined as the ratio of the true mass thickness (*A_j_ ≡ ρ_j_t_j_*) to the square root of the Cramer Rao Lower Bound (CRLB) on variance,

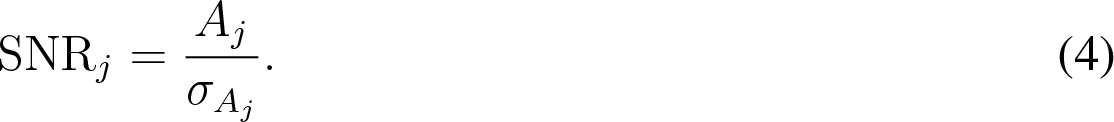

The CRLB was found from the Fisher information *F* using the known relation 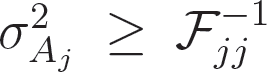.^27^ The signal noise model, which was energy-weighted compound Poisson, was approximated as a Gaussian with mean and variance matching the first two moments of the true distribution.^3^ The mean of each measurement is simply *λ_i_* as in Eq. 1, and the corresponding variance 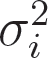 is

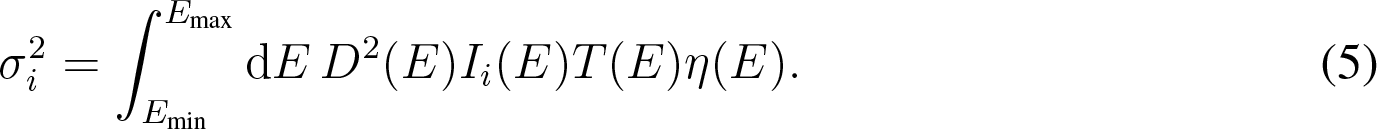

Thus, each DE acquisition (*i* = 1, 2) yields a Fisher information matrix with elements in terms of *λ_i_* and 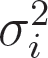 and their partial derivatives with respect to the true mass thicknesses *A*,

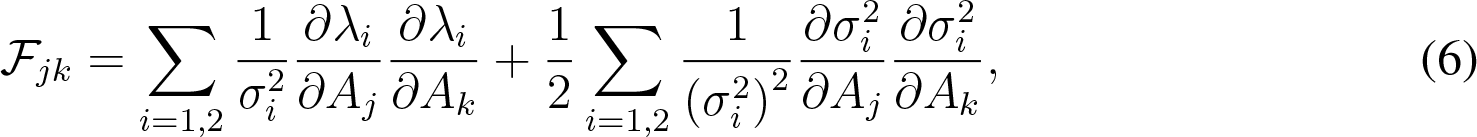

where *i* is the spectral index and *j, k* are the basis material indices.^21^

*2.3 Model 2: fan-beam CT of an anthropomorphic phantom*

To assess whether MV-kV DE-CT may provide advantages in clinical imaging scenarios, the signal detection framework was extended from the single-line model to a fan-beam CT geometry with 1200 views, 800 detector channels, and a fan angle of 47*^◦^*. A single 360*^◦^* rotation was simulated for each acquisition. Beam transmission through a computational anthropomorphic phantom, the extended cardiac torso (XCAT), was calculated.^28^ The phantom had dimensions of 512*×*512 with 1 mm^2^ pixels. Pathlengths through each pixel were determined using Siddon’s algorithm for the exact radiological path through a CT array.^29^ Since MeV photons are generally more penetrating than keV photons, the effect of high attenuation was considered by imaging the pelvis region with and without a surgical-grade stainless steel hip replacement (Fig. 3).^30^

**Fig 3.**
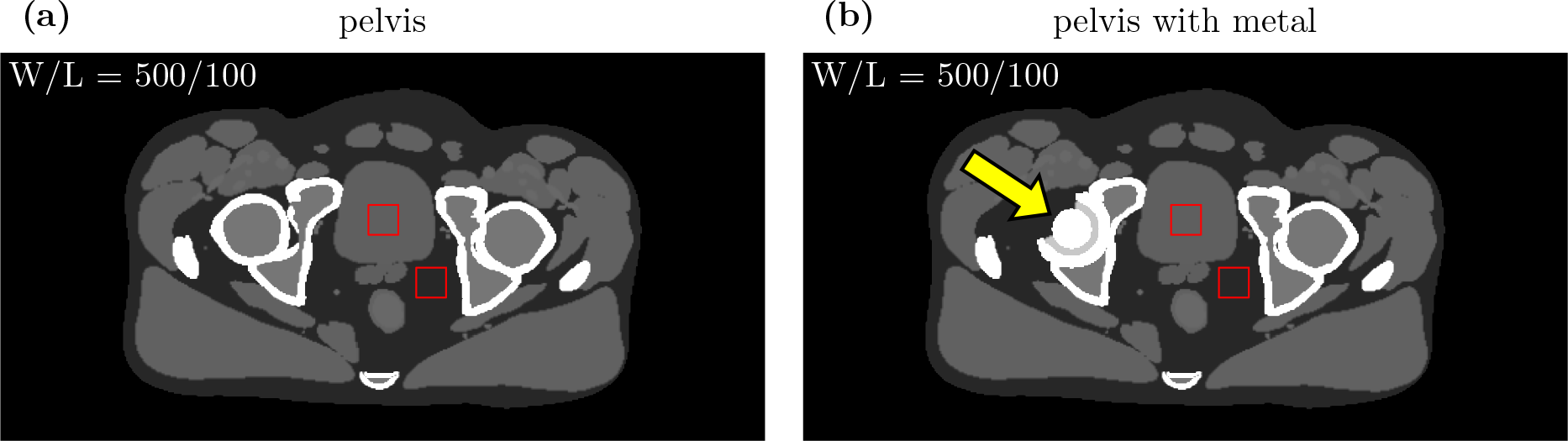
The two computational phantoms imaged (noiseless 80 keV monoenergetic images). The stainless steel hip replacement is indicated by the arrow in panel (b). CNR was computed using the delineated ROIs, and RMSE was evaluated within the phantom.

The total dose of each DE-CT acquisition was set to 10 mGy. Sinograms were generated for two spectral pairs, “MV-kV” (9 mGy detunedMV and 1 mGy 80kV) and “kV-kV” (5 mGy 140kV and 5 mGy 80kV). Each sinogram pair was decomposed into ICRU tissue and bone basis materials using a Gauss-Newton algorithm.^31^ While basis material decomposition may occur in either the sinogram or image domain, the sinogram-domain decomposition was chosen, because it has the advantage of ameliorating beam hardening artifacts. The basis material sinograms were then reconstructed into basis material images (BMIs) using fan-beam filtered back projection (FFBP) including a general sinc window with cutoff frequency at 80% of the Nyquist frequency.^32^ These BMIs correspond to the densities of the materials *ρ_i_*. The reconstructed images had a matrix size of 512*×*512 and field-of-view of 50 cm. Virtual monoenergetic images (VMIs) were generated at various energies *E*_0_ as a linear combination of the BMIs,

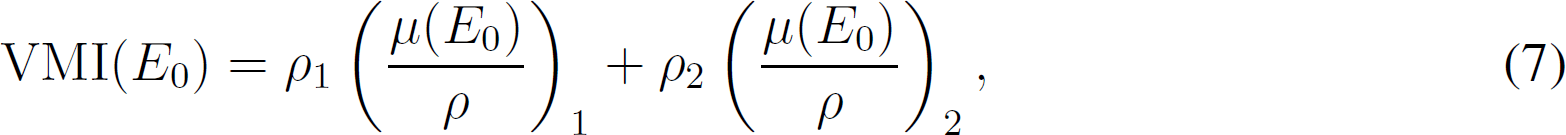

where (*µ*(*E*_0_)*/ρ*)_j_ is the known mass attenuation coefficient of basis material *j* at the energy *E*_0_. For comparison, three 10 mGy single-energy (SE) acquisitions were also generated (80kV, 120kV, and 140kV) and reconstructed using the same FFBP algorithm.

To evaluate contrast, CNR was computed in each SE-CT image and DE-CT VMI using measurements from the ROIs delineated in Fig. 3. CNR was defined as

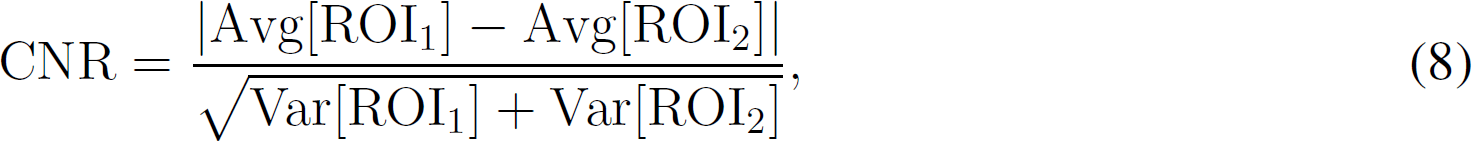

where ROI_1_ is the signal and ROI_2_ is the background.

To evaluate accuracy, the VMIs were registered to the input phantom, and root-mean-square-bone thickness [cm] error (RMSE) was computed relative to the monoenergetic ground truth,

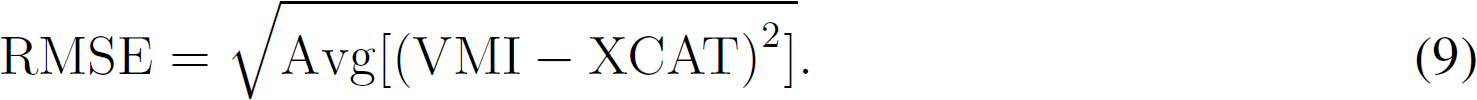

Note that SE-CT measurements are not energy-dependent, whereas CNR, RMSE, VMI, and XCAT include implicit energy dependence from each monoenergetic evaluation.

## 3 Results

### 3.1 Model 1: single-line integral

To identify the most promising spectral pairs, Fig. 4 and 5 present heatmaps of peak tissue and bone SNR, respectively, for each spectral combination and the ten bone thicknesses. Peak SNR was found from the curve of SNR as a function of dose allocation *r*. For both basis materials, the 140kV-80kV pair yields the highest SNRs overall. The detunedMV-80kV pair yields the highest SNRs of the MV-kV pairs. These two spectral pairs will be the focus of further analysis (“kV-kV” and “MV-kV,” respectively). Tissue SNR is maximized for both pairs with 1 cm bone, and bone SNR is maximized for kV-kV at 4 cm bone and MV-kV at 6 cm bone.

**Fig 4.**
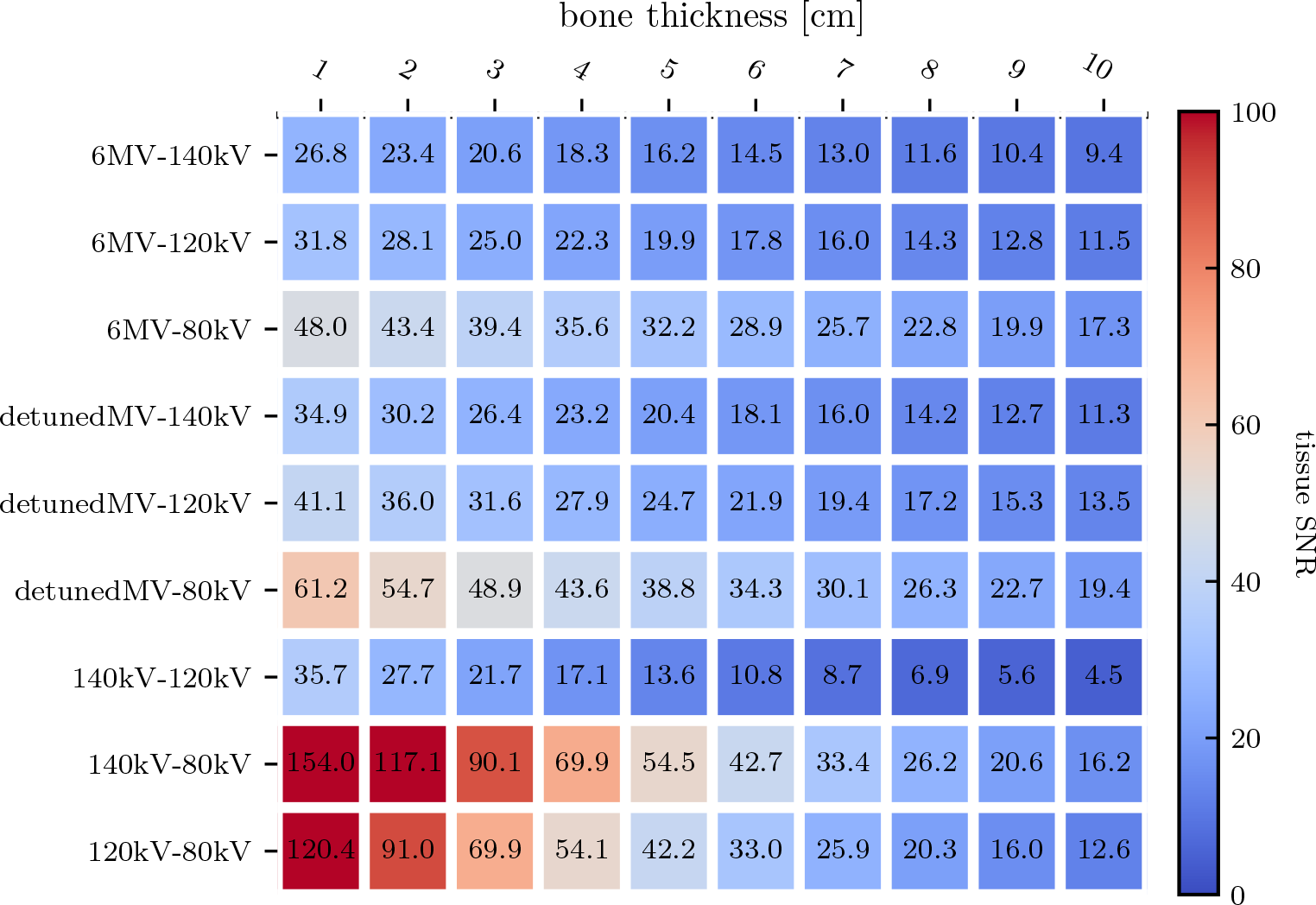
Heatmap of peak tissue SNR as a function of bone thickness for all dual-energy spectral combinations.

**Fig 5.**
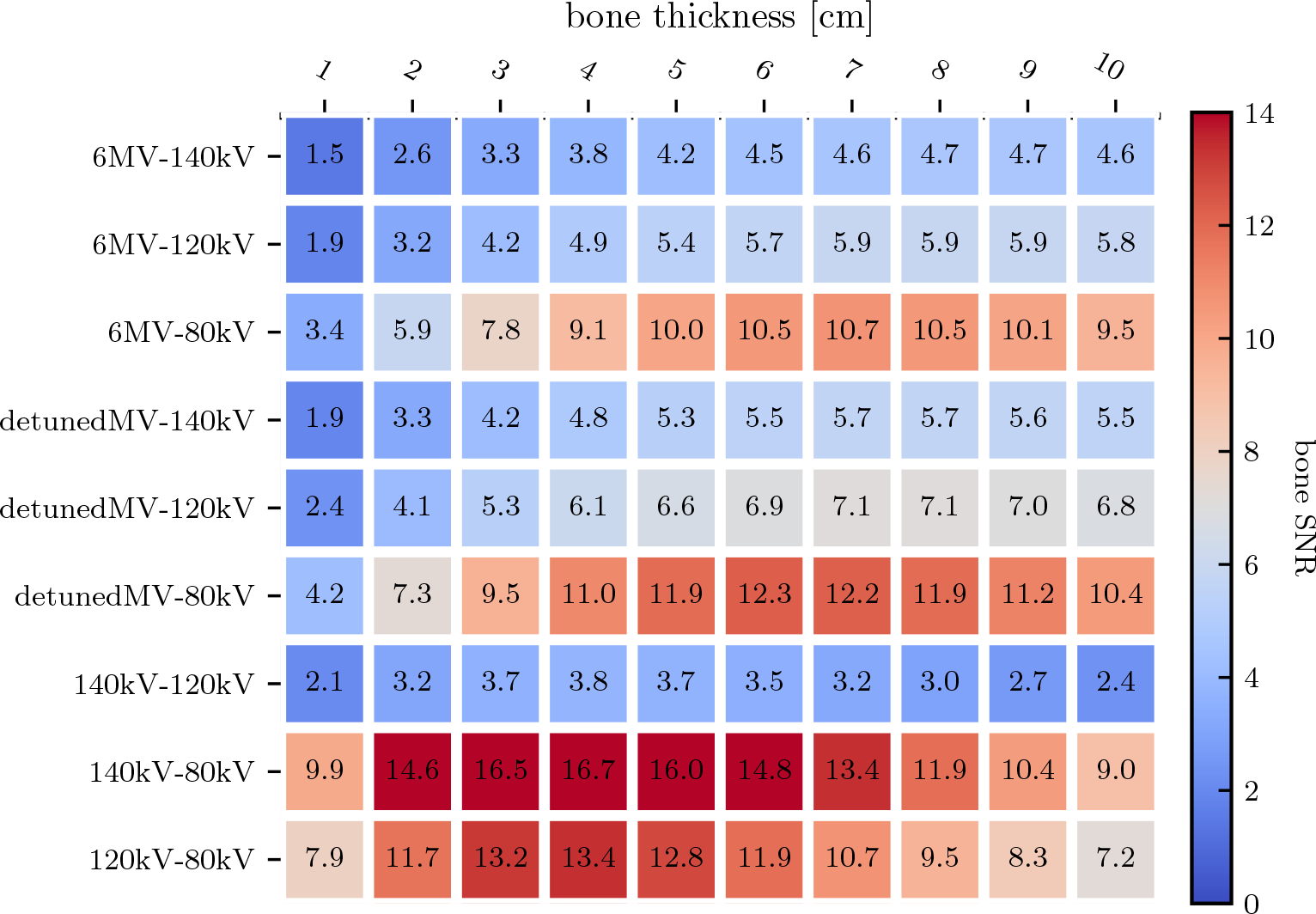
Heatmap of peak bone SNR as a function of bone thickness for all dual-energy spectral combinations.

Looking at optimal spectral dose distribution, Fig. 6 shows tissue basis material SNR as a function of high-energy dose allocation *r* for three different bone thicknesses. Based on the peaks in the two heatmaps, bone thicknesses of 1, 4, and 6 cm were chosen. Table 1 lists the coordinates of the peak basis material SNR for all spectral combinations with 1 cm bone. The MV-kV curve is skewed toward allocating a greater proportaion of dose to the MV spectrum, peaking at *r* = 0.92, 0.83, and 0.75 for 1, 4, and 6 cm, respectively. The kV-kV curve favors a more equal dose distribution, peaking at *r* = 0.51, 0.44, 0.40. As bone thickness increases, SNR is maximized by increasing the dose allocated to the low-energy spectrum.

**Fig 6.**
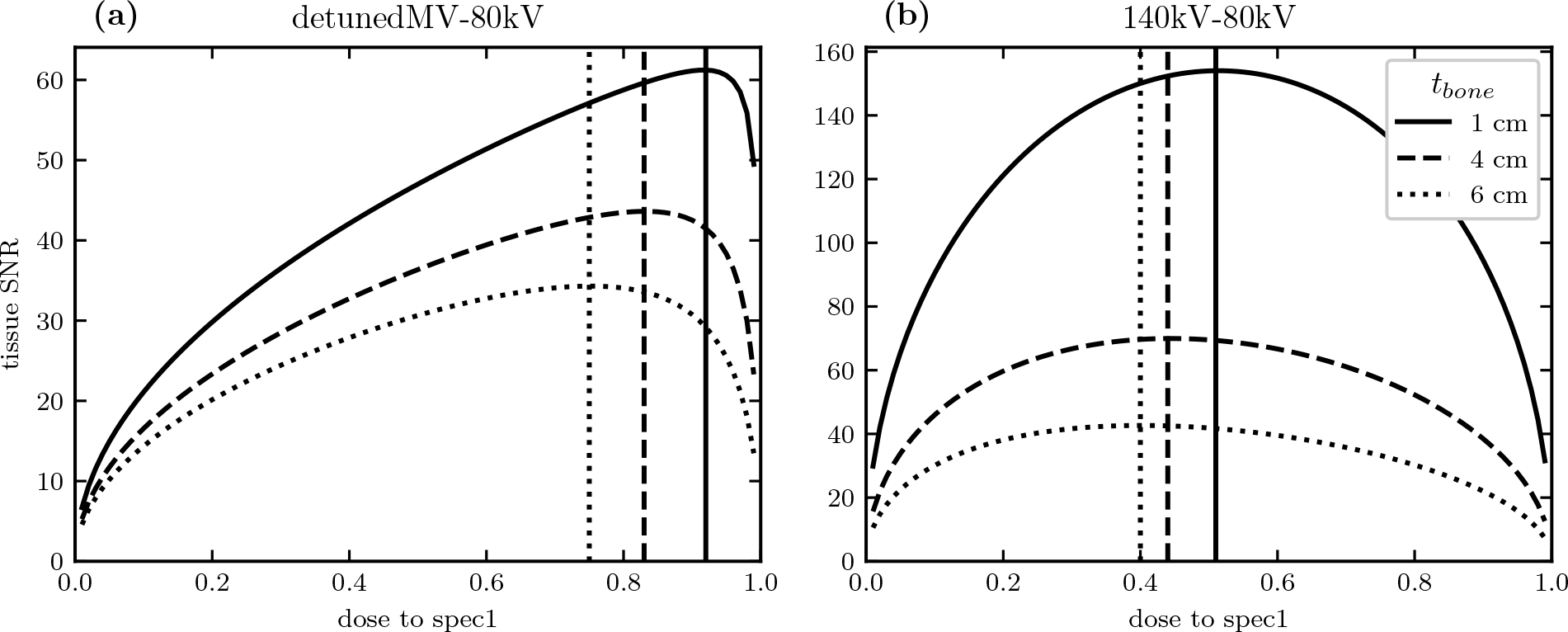
With 1, 4, or 6 cm bone thickness, curve of bone SNR as a function of dose allocated to the high-energy spectrum for the detunedMV-80kV (a) and 140kV-80kV (b) spectral pairs.

**Table 1.** Peak coordinates (*r*_max_, SNR_max_) as a function of dose allocation for both basis materials and all spectral pairs with a bone thickness of 1 cm.

| Spectral pair | Tissue |  | Bone |  |
| --- | --- | --- | --- | --- |
| | $r_{\max}$ | $\text{SNR}(r_{\max})$ | $r_{\max}$ | $\text{SNR}(r_{\max})$ |
| 6MV-80kV | 0.93 | 47.99 | 0.89 | 3.36 |
| 6MV-120kV | 0.93 | 31.79 | 0.91 | 1.86 |
| 6MV-140kV | 0.93 | 26.79 | 0.91 | 1.49 |
| detunedMV-80kV | 0.92 | 61.23 | 0.87 | 4.24 |
| detunedMV-120kV | 0.92 | 41.11 | 0.89 | 2.39 |
| detunedMV-140kV | 0.91 | 34.88 | 0.89 | 1.93 |
| 140kV-80kV | 0.51 | 154.02 | 0.45 | 9.89 |
| 140kV-120kV | 0.51 | 35.72 | 0.50 | 2.09 |
| 120kV-80kV | 0.50 | 120.38 | 0.45 | 7.94 |

To assess the effect of increasing object attenuation, Fig. 7 shows peak SNR as a function of bone thickness for the MV-kV and kV-kV pairs. At low bone thicknesses, kV-kV SNR is higher than MV-kV SNR for both basis materials. The tissue SNR monotonically decreases with bone thickness, while the bone SNR reaches a maximum at a thickness of 4 cm (kV-kV) or 6 cm (MV-kV). As bone thickness increases, the difference between the two curves decreases, and at 8 cm bone, the MV-kV curves intersect the kV-kV curves and begin to yield higher SNRs. This is due to the more rapid SNR drop-off of kV-kV imaging at high bone thickness.

**Fig 7.**
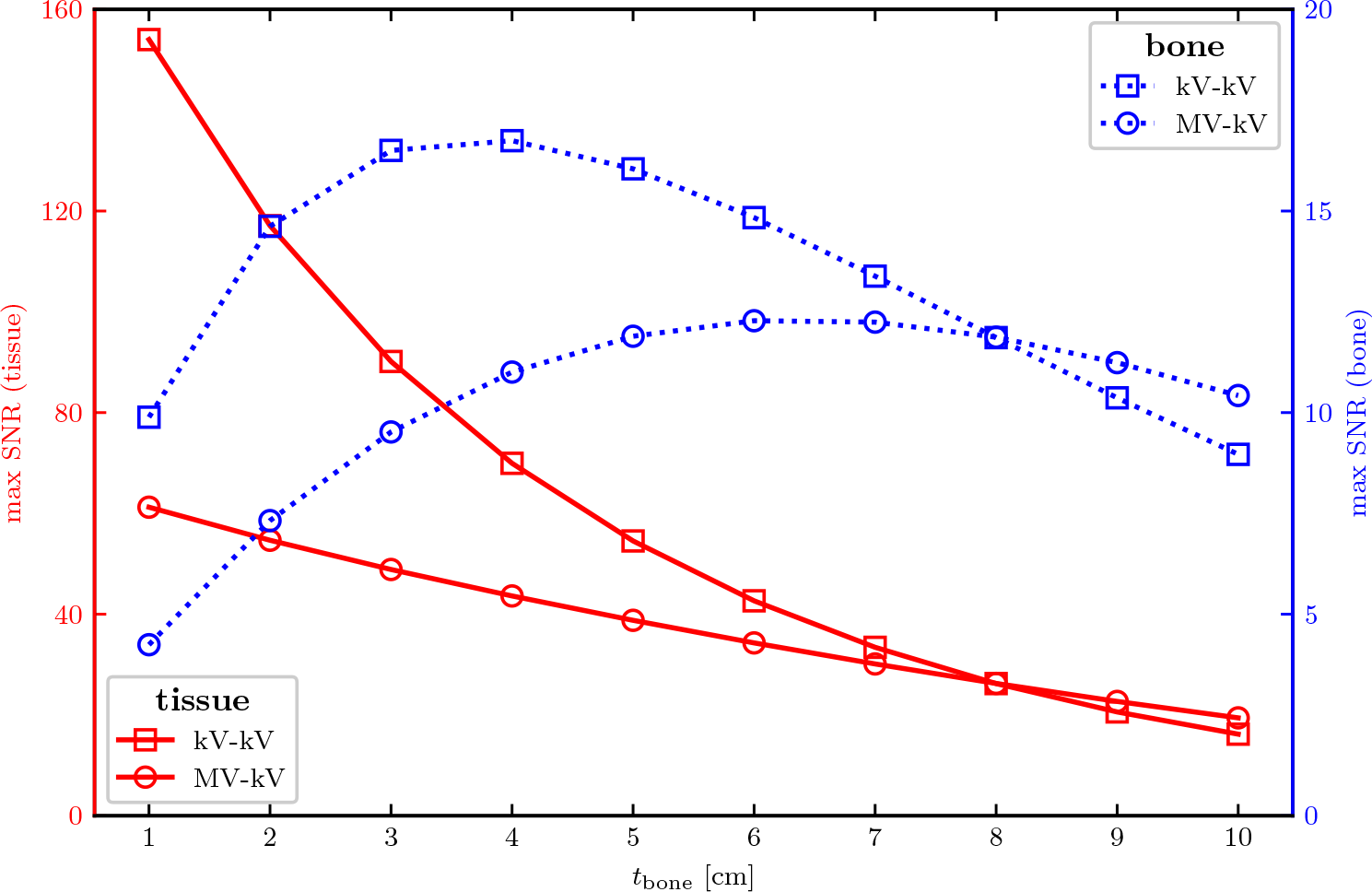
Maximum SNR for tissue (solid lines, left scale) and bone (dashed lines, right scale) for the 140kV-80kV (kV-kV, square marker) and detunedMV-80kV (MV-kV, circle marker) spectral combinations as a function of bone thickness.

### 3.2 Model 2: fan-beam CT

To quantify image quality in the simulated CT images, Fig. 8 shows CNR in the VMIs of the pelvis phantom with and without metal. CNR was measured as a function of VMI energy, and horizontal lines were drawn at the fixed CNR for each kV SE-CT image. These represent thresholds above which DE-CT may yield improved image quality for a given imaging task. For the pelvis without metal, this threshold is 2.73 (140kV), which is surpassed by both kV-kV and MV-kV DE-CT at 60 keV. The kV-kV CNR continues to increase with higher VMI energies, whereas the MV-kV CNR drops below the SE-CT threshold above 70 keV. With metal, all CNRs are considerably lower, and the SE-CT threshold is 0.30 (140kV). This is surpassed by kV-kV DE-CT above 100 keV and by MV-kV DE-CT above 130 keV. At higher VMI energies (*>*160 keV), MV-kV exceeds kV-kV DE-CT and yields the highest CNRs achieved by any CT acquisition evaluated. MV-kV CNR increases continually with VMI energy and begins to plateau above 300 keV.

**Fig 8.**
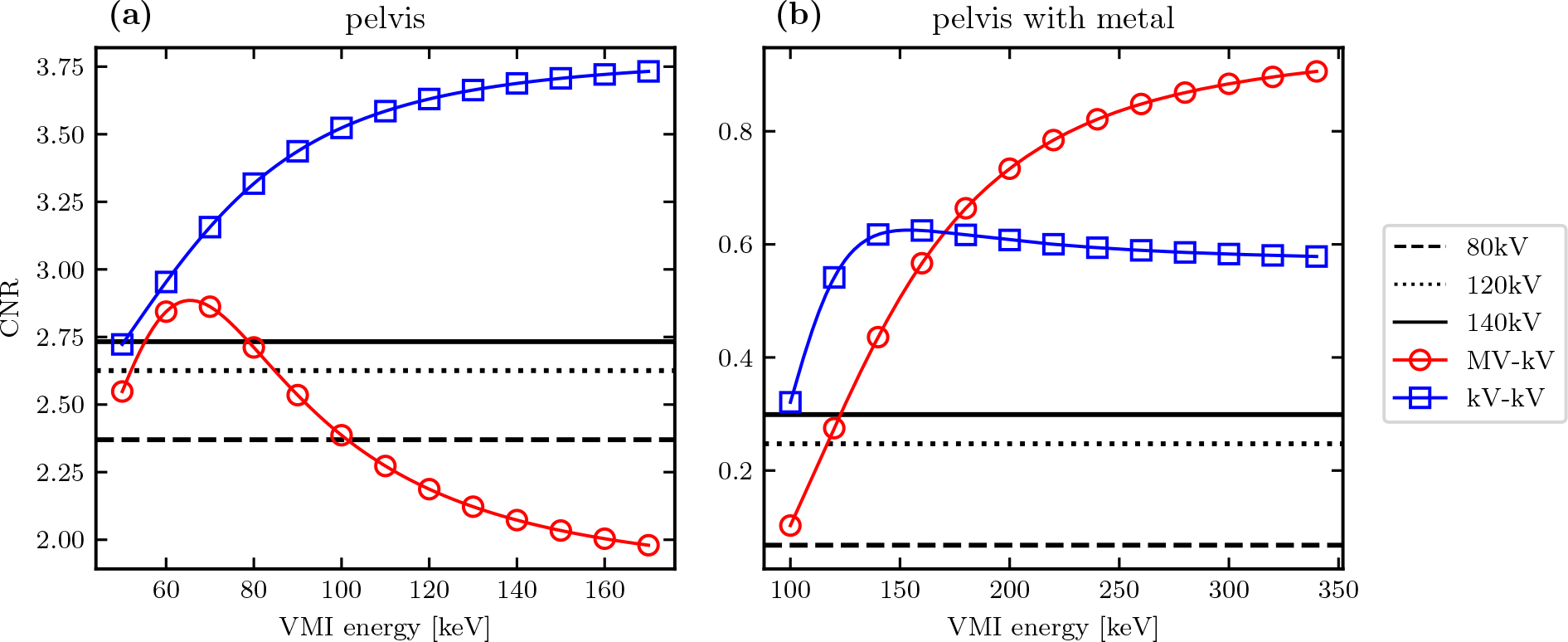
CNR in the detunedMV-80kV (MV-kV) and 140kV-80kV (kV-kV) VMIs as a function of energy, with horizontal lines at the CNR of each kV single-energy CT acquisition.

To quantify accuracy of the simulated CT images relative to the known ground truth, Fig. 9 shows RMSE for each DE-CT VMI. Generally, RMSE appears to decrease with greater VMI energy. Depending on the imaging setup, this lower error may come at the cost of higher CNR (Fig. 8). For the pelvis without metal, the minimum RMSE is 38.5 HU for kV-kV and 45.8 HU for MV-kV DE-CT. The smallest error overall is achieved with kV-kV DE-CT, though the difference relative to MV-kV is small (+7.3 HU). With metal, the RMSE measurements are much larger, likely due to the overall higher noise and stainless steel material. The minimum RMSE is 202.3 HU for kV-kV and 120.9 HU for MV-kV DE-CT. In this case, MV-kV DE-CT yields the smaller minimum error with a difference of 81.4 HU.

**Fig 9.**
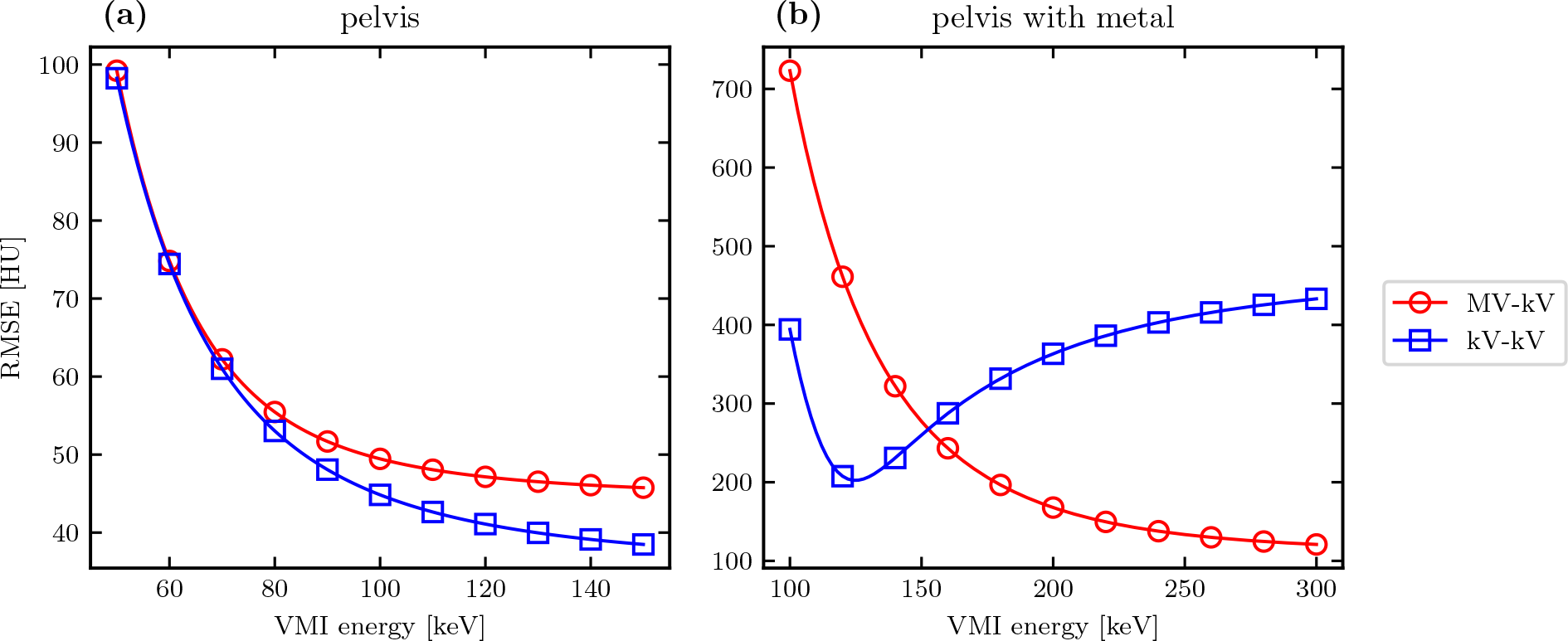
RMSE in the detunedMV-80kV (MV-kV) and 140kV-80kV (kV-kV) VMIs as a function of energy. RMSE was calculated relative to the monoenergetic ground truth XCAT.

For qualitative analysis, Fig. 10–13 show a sampling of SE-CTs, BMIs, and VMIs used for this analysis, providing complementary visual information for the measurements in Fig. 8 and 9. Figures 10 and 11 show the kV-kV and MV-kV DE-CT acquisitions of the pelvis without metal, and Fig. 12 and 13 show the same images with metal hip replacement. Qualitatively, in each kV CT, there is noticeable beam hardening in the pelvis and severe metal artifact streaking with the steel hip replacement. The detunedMV SE-CT suffers less apparent beam hardening, and although the metal hip increases noise, it does not cause as severe streaking. The kV-kV and MV-kV BMIs both show considerable streaking. In the MV-kV case, this is likely due to artifact contamination from the 80kV CT. Without metal, both MV-kV and kV-kV VMIs have visibly good contrast and no beam hardening. With metal, the 300 keV MV-kV VMI appears to have the least metal artifact contamination.

**Fig 10.**
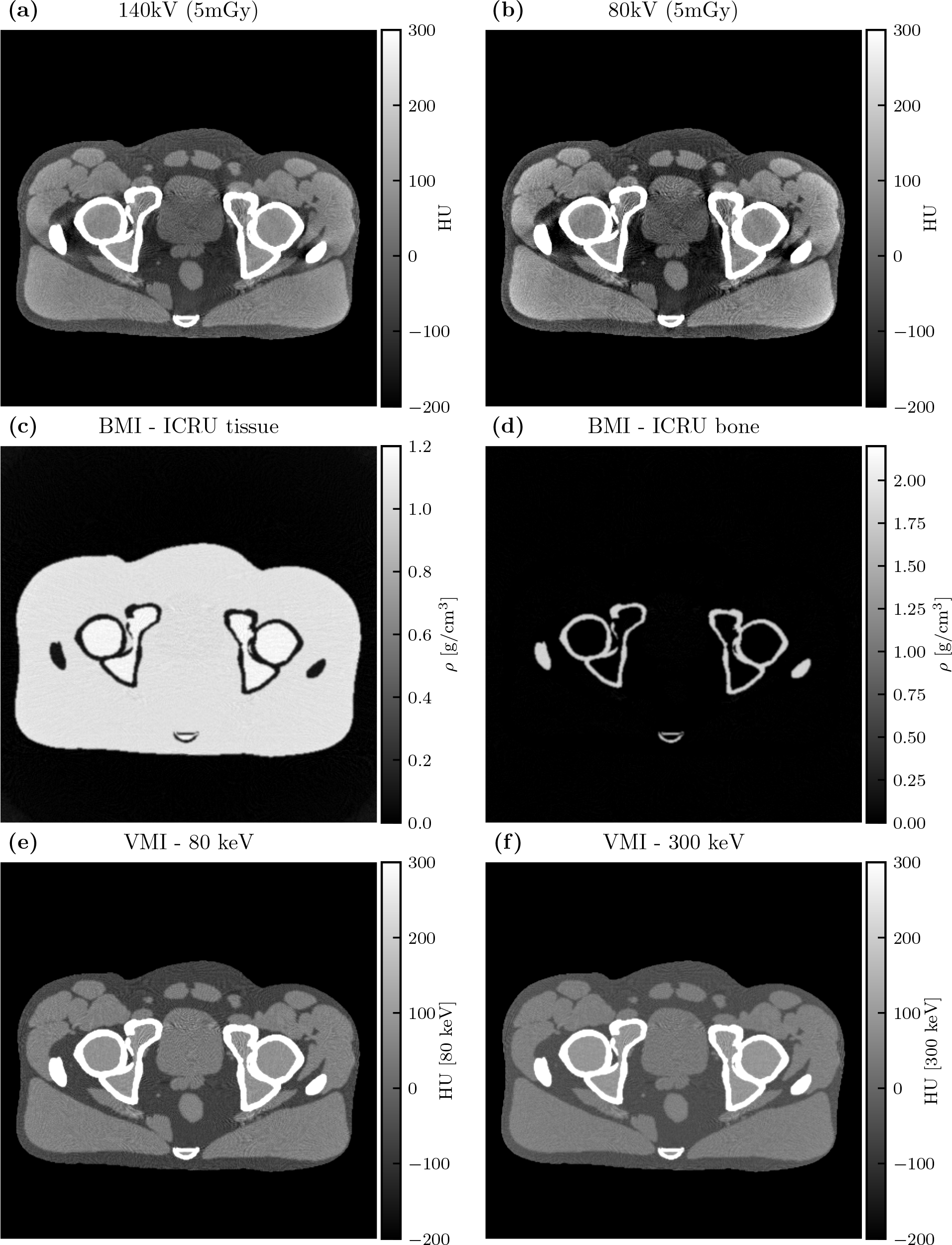
Single-energy CT images (a-b), basis material images (c-d), and VMIs (e-f) for the 140kV-80kV DE-CT acquisition.

**Fig 11.**
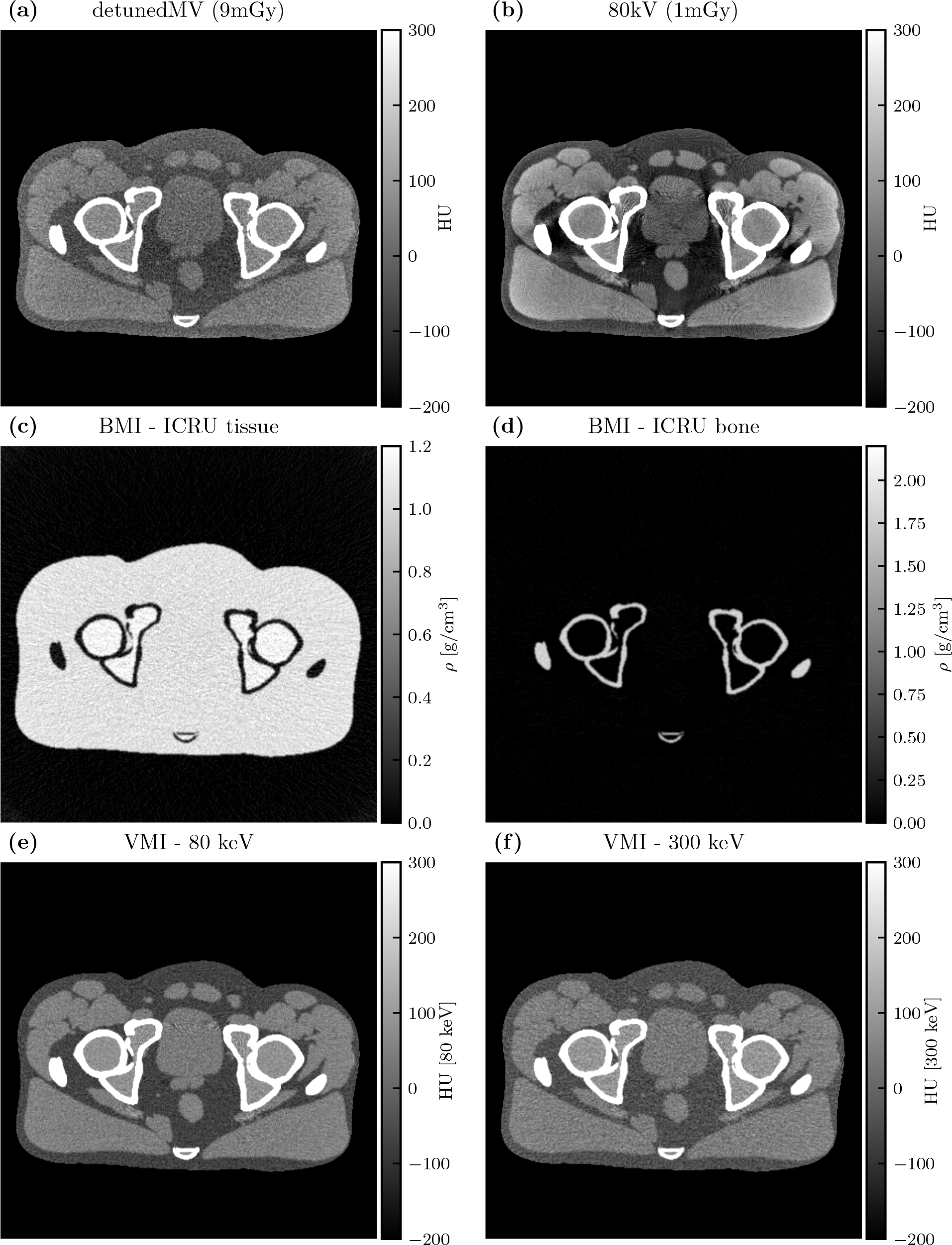
Single-energy images (a-b), basis material images (c-d), and VMIs (e-f) for the detunedMV-80kV DE-CT acquisition.

**Fig 12.**
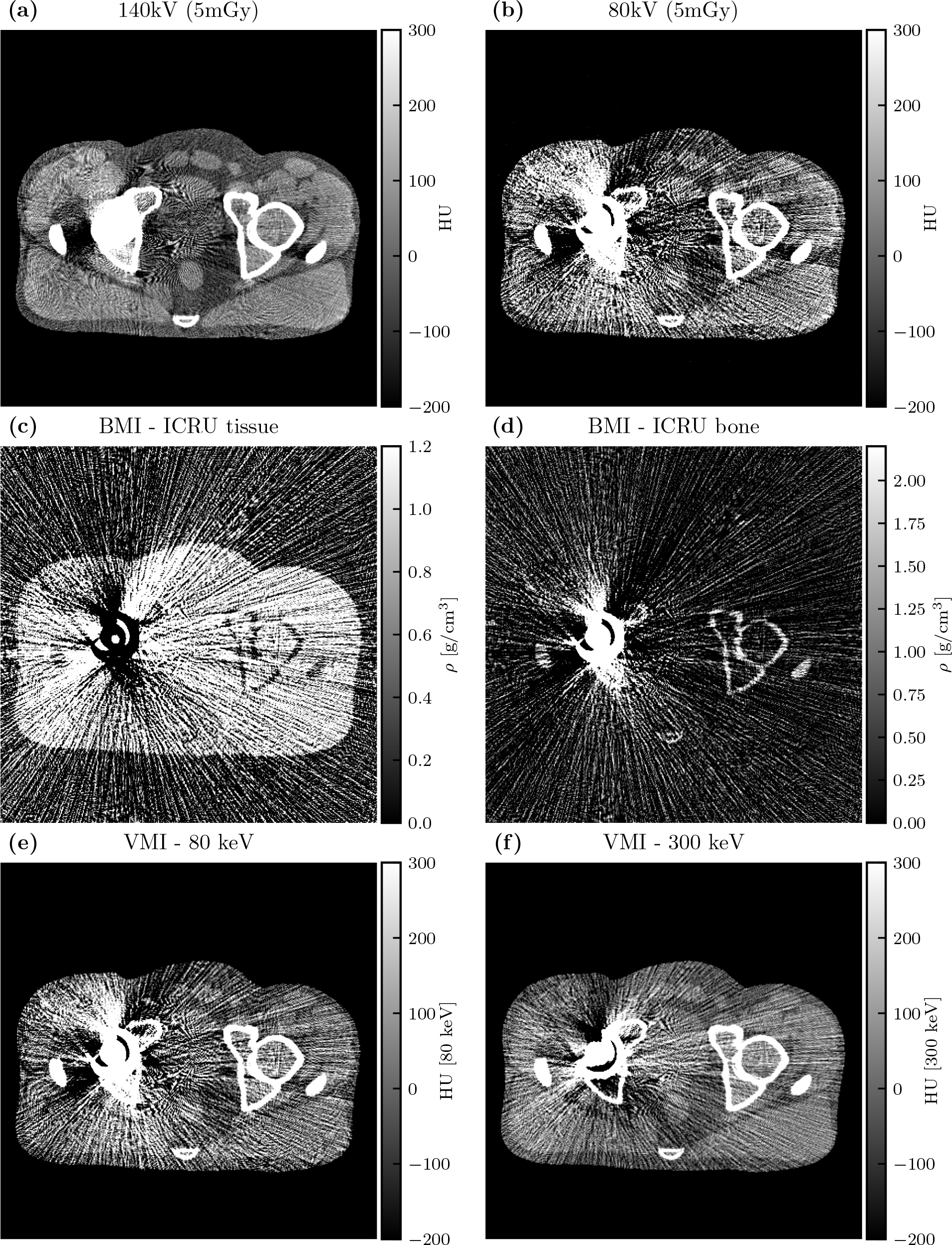
Single-energy images (a-b), basis material images (c-d), and VMIs (e-f) for the 140kV-80kV DE-CT acquisition with stainless steel hip replacement.

**Fig 13.**
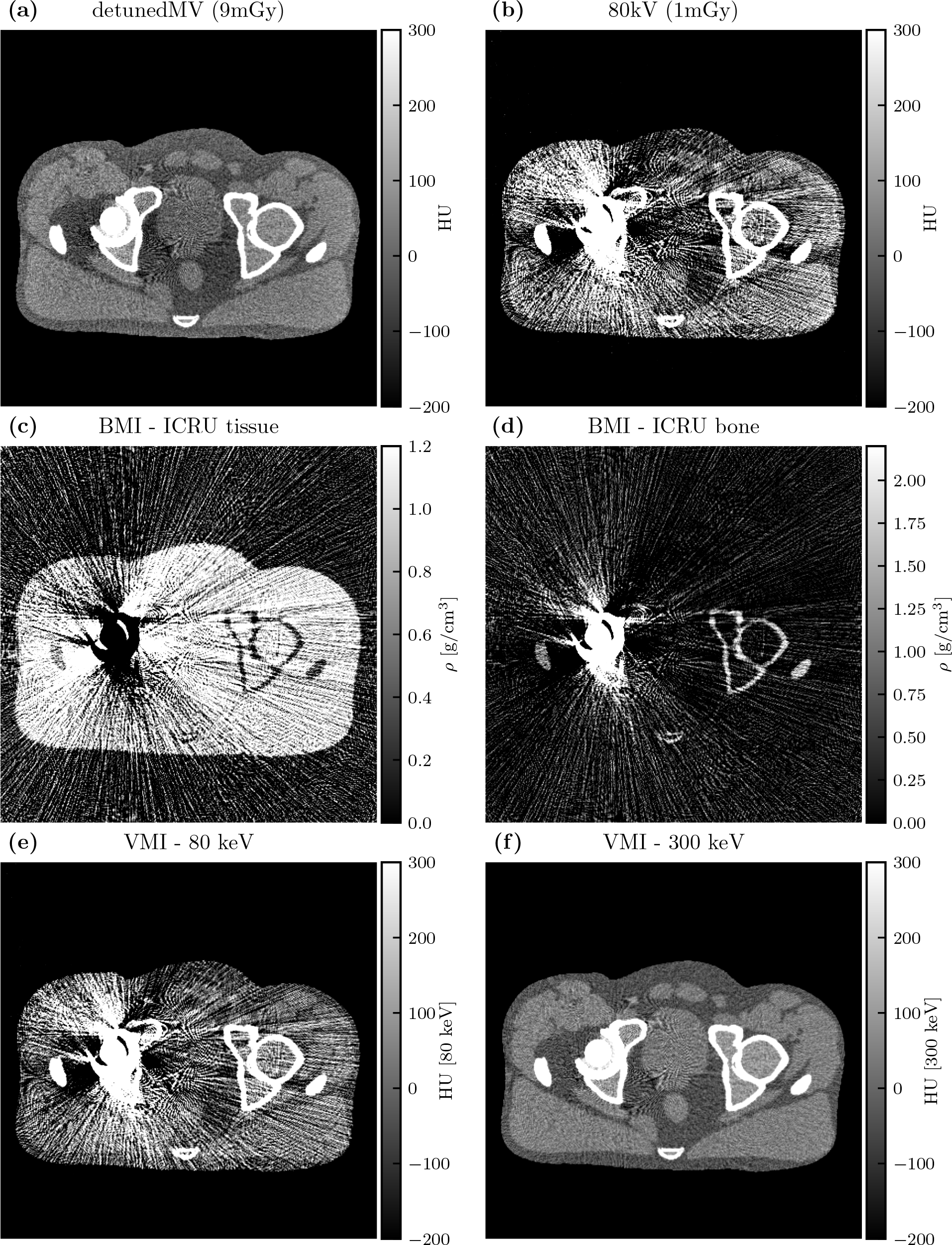
Single-energy images (a-b), basis material images (c-d), and VMIs (e-f) for the detunedMV-80kV DE-CT acquisition with stainless steel hip replacement.

## 4 Discussion

This work presents an evaluation of MV-kV DE-CT imaging, which could be realized using existing hardware in radiotherapy settings. We approached this task beginning with an estimation theory framework for calculating SNR along a single ray and then expanding to a simulated fan-beam CT acquisition of an anthropomorphic phantom.

Of the nine spectral pairs considered (six MV-kV and three kV-kV), the best basis material SNRs were found using the 140kV-80kV spectral pair. This is expected, as this spectral pair is commonly used for diagnostic DE-CT imaging, since it maximizes the energy separation between high- and low-energy spectra given the peak voltages conventionally available with modern x-ray tubes.^2^ Of the MV-kV DE pairs, the best SNRs were found using the detunedMV-80kV pair and the second-best with the 6MV-80kV pair. As the detuned beam has a lower effective energy than the treatment beam, it is expected to yield better image quality due to its higher detective efficiency and native contrast. However, the 6MV treatment beam has the potential to be used for acquiring images simultaneously with radiation therapy treatment, while the detuned beam could only be used for imaging before or after treatment.^10^ In this work, we chose to focus on the detuned beam. This choice provides a metric of the best achievable MV-kV CT image quality using beams currently available on therapy treatment systems, which is useful for an assessment of clinical viability.

We found that spectral dose distribution has a considerable effect on MV-kV image quality. As kV x-ray spectra have relatively similar effective energy, dose allocation may not be a typical consideration for diagnostic DE-CT. Single-ray basis material SNR peaked with approximately 90% dose allocated to the detunedMV spectrum or 50% dose to the 140kV spectrum when paired with the 80kV low-energy spectrum. The exact optimal dose allocation varied depending on the basis material and bone thickness. The asymmetric SNR-versus-dose allocation curve is a unique aspect of MV-kV DE-CT, likely owing to the greater dose deposition per photon with energy in the MeV range. In general, the optimal *r* was slightly higher for tissue SNR than for bone SNR. For both spectral pairs, *r* also decreased with increasing bone thickness. This relation was steeper for the MV-kV DE-CT pair. These results suggest spectral dose allocation is an important consideration for clinical implementation of MV-kV CT. Though past work assessing combined MV-kV image quality often draws comparisons with single-energy acquisitions, there is lack of a consideration of dose variations.^11, 14, 18–20^ A fair comparison of MV-kV DE-CT with single-energy CT should utilize dose-matched acquisitions with optimal spectral dose distributions depending on object composition. Our method provides one such way of carrying out this optimization.

For the fan-beam CT simulations, we chose the dose distribution between spectra using the single-ray approximations, with a 90:10 MV-kV distribution and 50:50 kV-kV distribution in each ray. However, across the many CT projection views and detector channels, each ray passes through a distinct section of the anatomy with its own bone and tissue thicknesses. This makes the task of optimal dose allocation more complex for CT than for a single ray. CT imaging traditionally uses bowtie filtration to reduce the dose allocated to more peripheral detector angles, producing more similar spectrum magnitudes after attenuation and, consequently, more uniform noise in each channel.^33^ Tube current modulation may be additionally implemented to modulate the dose allocated to each view angle, equalizing the noise in each projection.^34^ These methods further affect total dose and dose allocation across different object locations. The single-line model could be used to inform a joint bowtie filtration and tube current modulation algorithm that further optimizes image quality within DE-CT images while remaining dose neutral. Since the optimal dose allocation as a function of bone thickness varied more quickly for the detunedMV-80kV pair than it did for the diagnostic 140kV-80kV pair, MV-kV DE-CT image quality may especially benefit from such an algorithm, and it is worth considering for future applications.

As a function of bone thickness, tissue SNR monotonically decreased, and bone SNR peaked at 4 cm for the 140kV-80kV pair and 6 cm for the detunedMV-80kV pair. At low bone thicknesses, the SNR achieved with the kV-kV pair was higher than that with the MV-kV pair. This is more relevant for most imaging scenarios, especially at antero-posterior or posto-anterior CT view angles. However, at 8 cm bone thickness and greater, the MV-kV pair yields the highest SNRs. With the greater attenuation due to high bone thickness, the higher penetrability of the MeV photons becomes more beneficial as kV images begin to suffer from photon starvation. This effect has been utilized in other work for artifact correction around highly attenuating objects, namely metal implants.^17, 20^ Our findings corroborate this effect. For this reason, we explored DE-CT basis material image quality with and without metal implants in the XCAT phantom.

The simulated CT images show an advantage of MV-kV DE-CT over dose-matched single-energy kV images when imaging the pelvis. For comparison, we also simulated diagnostic kV-kV DE-CT images, although the technology for kV-kV DE-CT may not be readily available on radiotherapy systems. This is a unique practical advantage of MV-kV DE-CT. As expected, kV-kV DE-CT yielded the best image quality when imaging the pelvis. At energies up to 70 keV, both DE-CT VMIs yielded a similar CNR. At higher energies, kV-kV CNR continued to increase, but MV-kV CNR decreased below the kV CNR thresholds. Other work has similarly drawn comparisons between combined MV-kV images and single-energy kV images.^11, 14^ Li et al. observed that MV-kV VMIs of an image quality phantom can yield improved CNR relative to single-energy kV images with proper selection of virtual monoenergy.^14^ They found low monoenergy is preferable for low-density material inserts, and conversely, high monoenergy is preferable for high-density inserts. This trend matches our findings of CNR as a function of monoenergy in the pelvis with and without metal. Similarly, Yin et al. measured comparable or better contrast in aggregate MV-kV reconstructions compared to kV alone, depending on the material.^11^ It is relevant to note that these studies utilized a partial-angle acquisition technique and did not account for dose distribution, which is distinct from our method. In terms of accuracy compared to the ground truth phantom, we found that kV-kV RMSE was smaller than MV-kV RMSE at all energies, although the maximum difference with MV-kV VMIs was generally small. With the metal hip replacement, as expected, image quality suffered for all SE-CT acquisitions. The severity of metal artifacts was especially apparent in the kV images, and MV-kV VMIs yielded the best CNR and RMSE. Overall, these simulation results demonstrate the value of MV information for improving images beyond what is achievable with kV-only CT at the same dose.

This work was a simplified theoretical analysis of MV-kV DE-CT, and many limitations could be more realistically modeled in future work. Our compound Poisson noise model neglected x-ray scatter, patient motion, and electronic noise. A real CT acquisition will be affected by these complexities, and a DE-CT system using simultaneous acquisitions will also experience cross-scatter from the two beams. Though the simultaneous acquisition method introduces this additional scatter, it has the advantage of reducing motion artifacts relative to a sequential acquisition method. A Monte Carlo simulation could be additionally implemented to approximate both single-source scatter and dual-source cross-scatter, in order to better weigh the costs and benefits of each technique.^35, 36^ Other work has presented new methods for scatter reduction between MV and kV sources, which could also be considered.^37^ Though we did not model electronic noise, photon counting detectors are beginning to debut clinically, which have the advantage of thresholding out this noise. This could also be an avenue for future work. For image reconstruction, we implemented a standard filtered back-projection algorithm.^32^ More modern iterative and deep learning methods could be implemented, which may include more advanced noise reduction. Such algorithms likely especially benefit MV CT, since MV images tend to be noisier than kV images when dose-matched.

One potential extension of this work would be a more realistic differentiation of the two source-detector arrays. While traditional CT is acquired with a full field-of-view, some work has considered MV images that are truncated by multi-leaf collimators (MLCs).^10^ Future work could simulate or acquire MLC leakage images, especially with a simulated dose plan that opens the MLCs around the tumor. These images would use a higher incident flux, and attenuation through the MLCs would be calculated. Such images may suffer from a limited field-of-view and ring artifacts, for which corrections should be explored. Similarly, different detector materials and geometries should be considered for the MV and kV systems. In this work, we assumed the same detector was used for all acquisitions, with a fan shape and a fixed number of channels. This matched detector acquisition allows for material decomposition in the sinogram domain, which has the advantage of alleviating beam hardening artifacts. However, in current clinical settings, it is likely that the kV source would have a larger fan angle and more detector channels. The different sinogram properties will result in different reconstructed image qualities and necessitate implementation of an image-domain material decomposition algorithm, which may affect the resulting image quality. Nevertheless, the results of this initial investigation provide sufficient motivation for future studies with more faithful geometric modeling.

## 5 Conclusion

This work presents an analysis of MV-kV DE-CT imaging. We estimate that basis material SNR is maximized with 90% dose allocated to the MV spectrum. For bone thicknesses greater than 8 cm, SNR is maximized using MV-kV DE-CT. In a simulated pelvis scan, we find that MV-kV VMIs produce higher CNR than single-energy kV CT images. With a metal hip replacement, MV-kV VMIs can produce higher CNR and lower RMSE than diagnostic kV-kV VMIs. These results affirm the clinical utility of MV-kV DE-CT and more robustly quantify the parameters for optimal implementation.

### Disclosures

Patrick La Rivière has in the past received research funding from Accuray Inc., Canon Medical Research USA, Inc., and Toshiba Medical Research USA, Inc. He is on the Scientific Advisory Board and holds stock options in Metritrack, Inc, an ultrasound-guidance company.

### Code, Data, and Materials Availability

The code used to generate the data in this manuscript can be freely accessed through GitHub at https://github.com/gjadick/dex-single-ray for the single-line integral toy model and https://github.com/gjadick/dex-ct-sim for the fan-beam CT simulation and basis material decomposition.

## Acknowledgments

We would like to thank Julian Bertini for his contributions to the single-line integral toy model, Phillip Vargas for insightful discussions, and Accuray, Inc. for early funding of this work.

**Giavanna Jadick** is a Ph.D. student in the Graduate Program in Medical Physics at the University of Chicago. She received her BS degree in physics from Duke University in 2020. Her research focuses on multi-energy computed tomography imaging. She is a student member of SPIE.

**Geneva Schlafly** is a Ph.D. student in the Graduate Program in Medical Physics at the University of Chicago. She received her BS degree in mathematics from the University of California, Santa Barbara in 2020. Her research is in tomography techniques for polarized light microscopy. She is Vice President of the SPIE University of Chicago Student Chapter.

**Patrick J. La Rivière** is a professor in the Department of Radiology and the Committee on Medical Physics at the University of Chicago. He received his AB degree in physics from Harvard University and his PhD from the Graduate Program in Medical Physics at the University of Chicago. His research interests include tomographic reconstruction in computed tomography, x-ray fluorescence computed tomography, and computational microscopy. He is a fellow of SPIE.

